# Adapting the UK Biobank brain imaging protocol and analysis pipeline for the C-MORE multi-organ study of COVID-19 survivors

**DOI:** 10.1101/2021.05.19.21257316

**Authors:** Ludovica Griffanti, Betty Raman, Fidel Alfaro-Almagro, Nicola Filippini, Mark Philip Cassar, Fintan Sheerin, Thomas W Okell, Flora A Kennedy McConnell, Michael A Chappell, Chaoyue Wang, Christoph Arthofer, Frederik J Lange, Jesper Andersson, Clare E Mackay, Elizabeth Tunnicliffe, Matthew Rowland, Stefan Neubauer, Karla L Miller, Peter Jezzard, Stephen M Smith

## Abstract

SARS-CoV-2 infection has been shown to damage multiple organs, including the brain. Multiorgan MRI can provide further insight on the repercussions of COVID-19 on organ health but requires a balance between richness and quality of data acquisition and total scan duration. We adapted the UK Biobank brain MRI protocol to produce high-quality images while being suitable as part of a post-COVID-19 multiorgan MRI exam. The analysis pipeline, also adapted from UK Biobank, includes new imaging-derived phenotypes (IDPs) designed to assess the effects of COVID-19. A first application of the protocol and pipeline was performed in 51 COVID-19 patients post-hospital discharge and 25 controls participating in the Oxford C-MORE study.

The protocol acquires high resolution T_1_, T_2_-FLAIR, diffusion weighted images, susceptibility weighted images, and arterial spin labelling data in 17 minutes. The automated imaging pipeline derives 1575 IDPs, assessing brain anatomy (including olfactory bulb volume and intensity) and tissue perfusion, hyperintensities, diffusivity, and susceptibility. In the C-MORE data, these quantitative measures were consistent with clinical radiology reports. Our exploratory analysis tentatively revealed that recovered COVID-19 patients had a decrease in frontal grey matter volumes, an increased burden of white matter hyperintensities, and reduced mean diffusivity in the total and normal appearing white matter in the posterior thalamic radiation and sagittal stratum, relative to controls. These differences were generally more prominent in patients who received organ support. Increased T_2_* in the thalamus was also observed in recovered COVID-19 patients, with a more prominent increase for non-critical patients.

This initial evidence of brain changes in COVID-19 survivors prompts the need for further investigations. Follow-up imaging in the C-MORE study is currently ongoing, and this protocol is now being used in large-scale studies. The pipeline is widely applicable and will contribute to new analyses to hopefully clarify the medium to long-term effects of COVID-19.

**Highlights:**

- UK Biobank brain MRI protocol and pipeline was adapted for multiorgan MRI of COVID-19
- High-quality brain MRI data from 5 modalities are acquired in 17 minutes
- Analysis pipeline derives 1575 IDPs of brain anatomy, perfusion, and microstructure
- Evidence of brain changes in COVID-19 survivors was found in the C-MORE study
- This MRI protocol is now being used in multiple large-scale studies on COVID-19

## INTRODUCTION

Since its outbreak in 2019, there has been a global effort to understand the impact of SARS-CoV-2 infection on organ health, both in the acute and medium-to-long term phases. Despite being predominantly a respiratory illness, emerging data suggest that damage to multiple organs is common, particularly among those with moderate to severe infections (Guan et al., 2020; Huang et al., 2020). A holistic approach to systematically assess the health of multiple vital organs could therefore be advantageous.

Recently we set out to evaluate the medium-term effects of COVID-19 on multiple vital organs (brain, lungs, heart, liver and kidneys) using magnetic resonance imaging (MRI) in the ‘Capturing MultiORgan Effects of COVID-19’ (C-MORE) study (Raman et al., 2021). A concise yet comprehensive protocol sensitive to the diverse range of COVID-19 manifestations was needed to ensure multiorgan coverage over a reasonable time frame. Incorporation of brain imaging was particularly vital in light of the neuroimaging findings observed in COVID-19 patients (Choi and Lee, 2020; Lu et al., 2020; Sawlani et al., 2021) and of the increasing number of studies suggesting a high prevalence of neurological symptoms, mental health abnormalities and cognitive impairment in survivors (Ellul et al., 2020; Mao et al., 2020; Taquet et al., 2021; Varatharaj et al., 2020). Reports of subclinical pathology including ischaemic and hemorrhagic events in patients recovering from moderate to severe infections have also raised concerns about the long-term neurological damage caused by COVID-19 (Miners et al., 2020).

In 2015, the UK Biobank (UKB) imaging study was launched with the primary aim of improving disease prevention, diagnosis and treatment through insights gained from high-quality imaging (Miller et al., 2016). As part of this effort, comprehensive imaging of the brain and other organs were planned in 100,000 patients of the original UKB cohort. To date, more than 40,000 patients have been scanned (as of May 2021). To facilitate high throughput image processing of brain images, an automated image analysis pipeline was also developed (Alfaro-Almagro et al., 2018), permitting rapid and reproducible image analysis of large datasets. The pipeline also extracts so-called imaging derived phenotypes (IDPs), quantitative measures that can be easily used and interpreted also by non-imaging experts. In view of this rich resource, the C-MORE study set out to align their post-COVID neuroimaging with the UKB protocol, however, there were two challenges to overcome. First, the standard 30-minute UKB brain protocol was too long to be directly incorporated into the 70-minute multiorgan protocol. Second, the existing UKB protocol and analysis pipeline was not customised to extract COVID-19 specific imaging markers of interest.

In this study, we sought to describe how we adapted the UKB brain MRI protocol to: 1) achieve an optimal scan time of under 20 minutes for incorporation into the multiorgan protocol; 2) augment its sensitivity to COVID-19 specific pathology 3) exploit its technical advances and enable future data comparison and merging. As an extension to the original study (Raman et al., 2021), the pipeline has been expanded to generate 1467 additional IDPs (cortical grey matter density measures, cortical thickness and area from FreeSurfer, olfactory bulb volume and intensity, mean diffusivity within the tracts’ normal appearing white matter, and perfusion metrics in the white matter, for a total of 1575 IDPs) and includes an improved estimate of quantitative susceptibility mapping of subcortical structures. We then assess the agreement between IDPs and radiology reports. Regarding the exploratory analyses on the impact of COVID-19 on the brain in the C-MORE dataset, we expanded the group comparison between patients and controls to include additional new IDPs, and investigated group differences based on disease severity and the presence or absence of anosmia.

## METHODS

### MRI acquisition protocol

Table 1 reports the details of the brain MRI sequences included in the multiorgan COVID-19 protocol, in comparison with the UKB protocol. Below we describe the rationale for the inclusion of these MRI sequences and, where applicable, deviations from the UKB protocol.

**Table 1.**
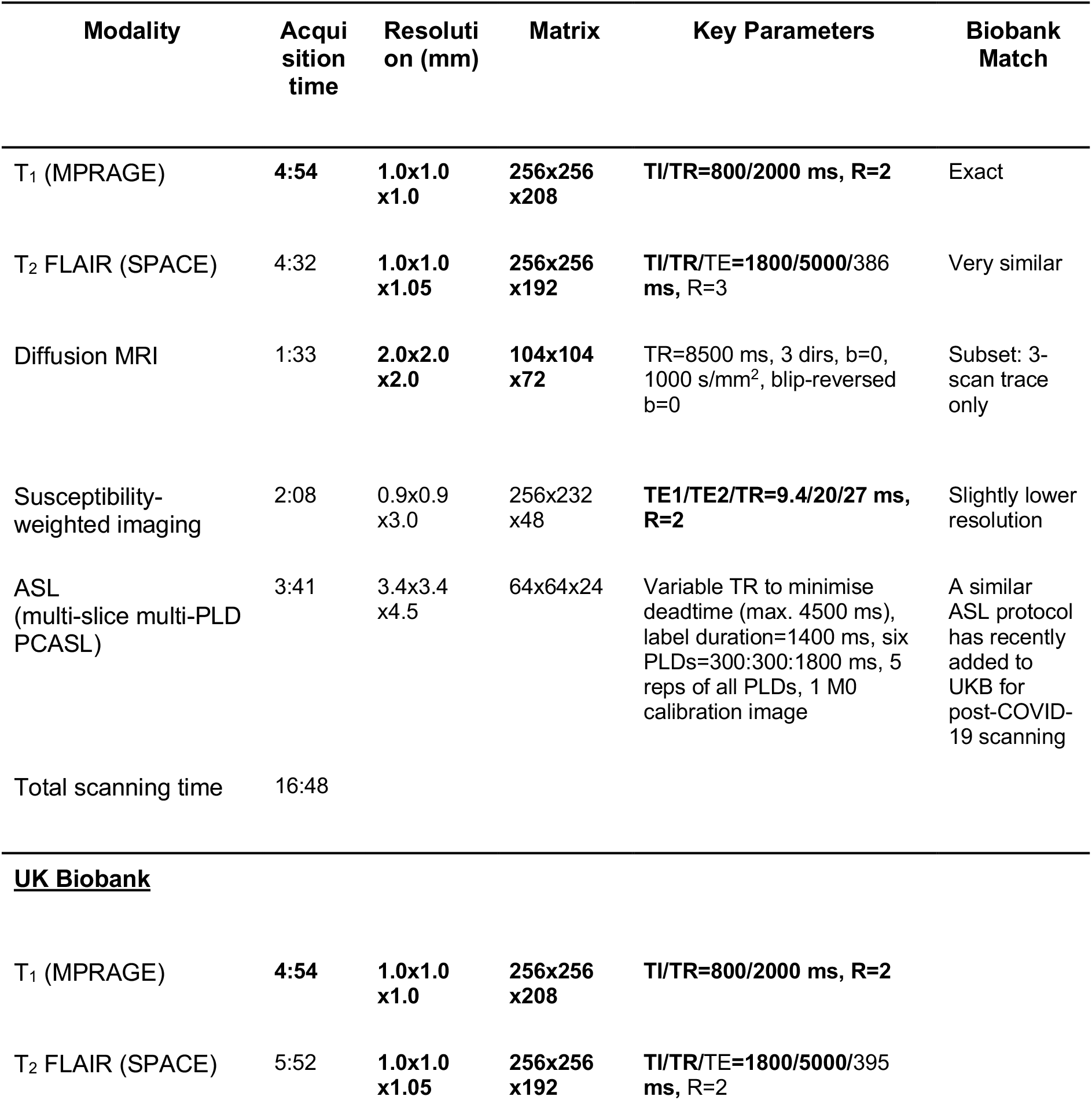
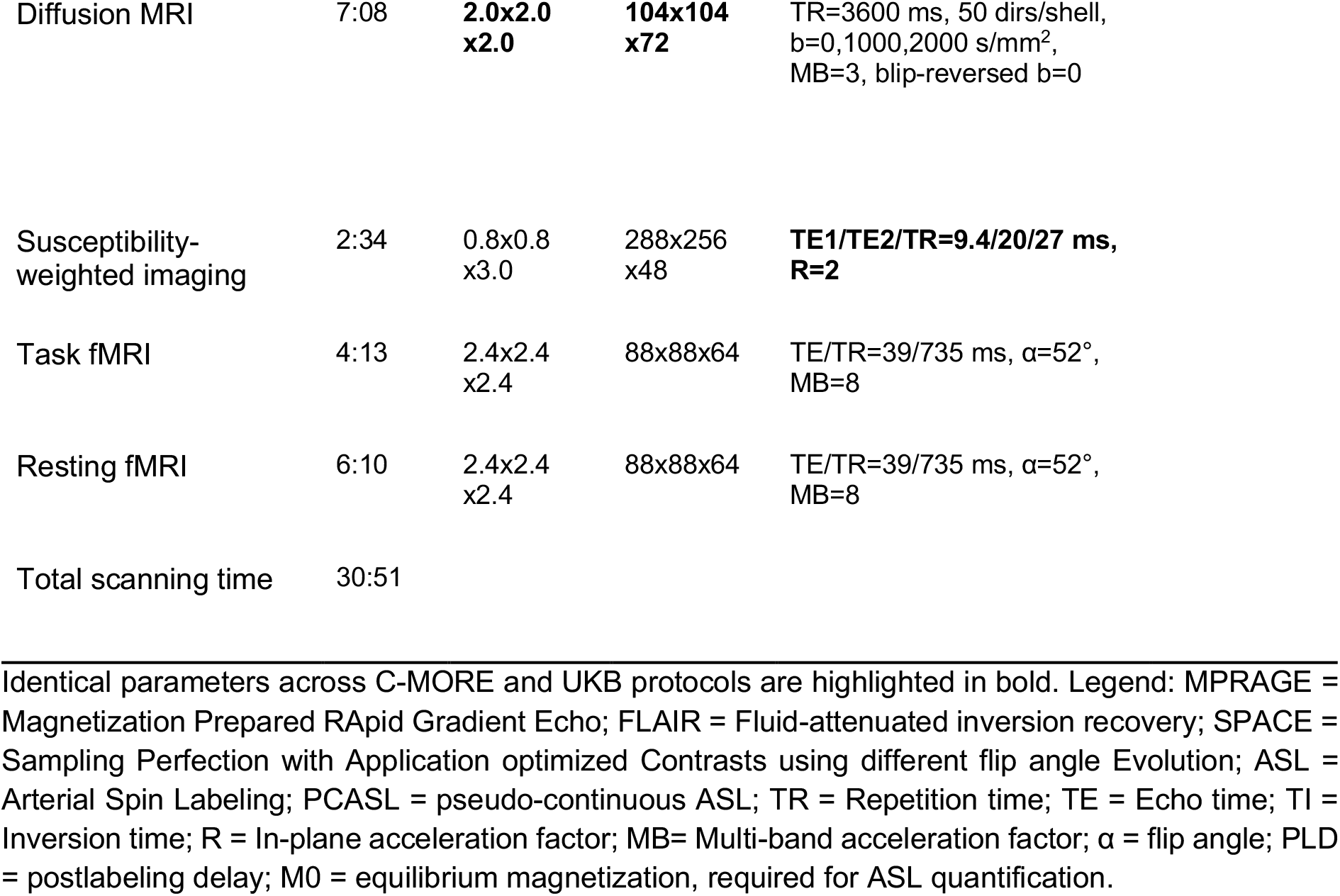
Brain MRI sequences of the COVID-19 multiorgan protocol: acquisition details and comparison with UKB protocol.

| Modality | Acquisition time | Resolution (mm) | Matrix | Key Parameters | Biobank Match |
| --- | --- | --- | --- | --- | --- |
| T<sub>1</sub> (MPRAGE) | 4:54 | 1.0x1.0x1.0 | 256x256x208 | TI/TR=800/2000 ms, R=2 | Exact |
| T<sub>2</sub> FLAIR (SPACE) | 4:32 | 1.0x1.0x1.05 | 256x256x192 | TI/TR/TE=1800/5000/386 ms, R=3 | Very similar |
| Diffusion MRI | 1:33 | 2.0x2.0x2.0 | 104x104x72 | TR=8500 ms, 3 dirs, b=0, 1000 s/mm<sup>2</sup>, blip-reversed b=0 | Subset: 3-scan trace only |
| Susceptibility-weighted imaging | 2:08 | 0.9x0.9x3.0 | 256x232x48 | TE1/TE2/TR=9.4/20/27 ms, R=2 | Slightly lower resolution |
| ASL (multi-slice multi-PLD PCASL) | 3:41 | 3.4x3.4x4.5 | 64x64x24 | Variable TR to minimise deadtime (max. 4500 ms), label duration=1400 ms, six PLDs=300:300:1800 ms, 5 reps of all PLDs, 1 M0 calibration image | A similar ASL protocol has recently added to UKB for post-COVID-19 scanning |
| Total scanning time | 16:48 |  |  |  |  |

UK Biobank
|  |  |  |  |  |
| --- | --- | --- | --- | --- |
| T<sub>1</sub> (MPRAGE) | 4:54 | 1.0x1.0x1.0 | 256x256x208 | TI/TR=800/2000 ms, R=2 |
| T<sub>2</sub> FLAIR (SPACE) | 5:52 | 1.0x1.0x1.05 | 256x256x192 | TI/TR/TE=1800/5000/395 ms, R=2 |
| Diffusion MRI | 7:08 | **2.0x2.0**<br>**x2.0** | **104x104**<br>**x72** | TR=3600 ms, 50 dirs/shell,<br>b=0,1000,2000 s/mm<sup>2</sup>,<br>MB=3, blip-reversed b=0 |
| Susceptibility-weighted imaging | 2:34 | 0.8x0.8<br>x3.0 | 288x256<br>x48 | **TE1/TE2/TR=9.4/20/27 ms,**<br>**R=2** |
| Task fMRI | 4:13 | 2.4x2.4<br>x2.4 | 88x88x64 | TE/TR=39/735 ms, $\alpha=52^\circ$,<br>MB=8 |
| Resting fMRI | 6:10 | 2.4x2.4<br>x2.4 | 88x88x64 | TE/TR=39/735 ms, $\alpha=52^\circ$,<br>MB=8 |
| Total scanning time | 30:51 |  |  |  |

Identical parameters across C-MORE and UKB protocols are highlighted in bold. Legend: MPRAGE = Magnetization Prepared Rapid Gradient Echo; FLAIR = Fluid-attenuated inversion recovery; SPACE = Sampling Perfection with Application optimized Contrasts using different flip angle Evolution; ASL = Arterial Spin Labeling; PCASL = pseudo-continuous ASL; TR = Repetition time; TE = Echo time; TI = Inversion time; R = In-plane acceleration factor; MB= Multi-band acceleration factor; $\alpha$ = flip angle; PLD = postlabeling delay; M0 = equilibrium magnetization, required for ASL quantification.

The scanner used in UKB is a standard Siemens Skyra 3T running VD13A, with a standard Siemens 32-channel RF receive head coil. The multiorgan COVID-19 protocol was setup on a Siemens Prisma 3T running VE11C, with a Siemens 20-channel head coil.

### Rationale for using specific brain MRI sequences in the COVID-19 multiorgan protocol

#### High-resolution T_1_-weighted image (T_1_)

This structural sequence is primarily used to study grey matter (GM) macroscopic anatomy in both cortical and subcortical brain regions. By exploiting differences in the interaction of water with surrounding tissues (tissue T_1_ relaxation times), this sequence provides strong contrast between grey and white matter. GM reductions have been widely associated with Alzheimer’s disease and age-related cognitive dysfunction (Pini et al., 2016). Emerging evidence shows that COVID-19 may exacerbate or even cause cognitive problems (Kremer et al., 2020; PHOSP-COVID Collaborative Group et al., 2021; Varatharaj et al., 2020), potentially contributing to a new wave of dementia and multi-morbidity in the future (Miners et al., 2020). A recent study (Lu et al., 2020), however, found GM increases in recovered hospitalised COVID-19 patients (MRI about 3 months from onset), relative to controls. High-resolution T_1_ is also critical for surface generation and calculations of cross-subject and cross-modality alignments, needed in order to process all other brain modalities and to perform voxel-wise analyses. This protocol was exactly matched to the one used in UKB.

#### T_2_-weighted Fluid Attenuated Inversion Recovery (T_2_-FLAIR)

This structural sequence is commonly used in clinical practice, for example to characterise white matter (WM) hyperintensities (WMH). In T_2_-weighted images, the contrast is dominated by signal decay from interactions between water molecules (T_2_ relaxation times) and image intensity is related to alterations to tissue compartments typically associated with pathology. While a recent review (Egbert et al., 2020) described WMHs as the most frequently reported brain abnormalities in adults with COVID-19 in the acute and subacute phases, other patterns of T_2_-FLAIR alterations have also been reported, including signal abnormalities in the medial temporal lobe and non-confluent multifocal WM hyperintense lesions with variable enhancement associated with hemorrhagic lesions (Kremer et al., 2020). Loss of smell, a characteristic symptom of COVID-19, has been linked with abnormal olfactory bulb T_2_-FLAIR signal (Lin et al., 2020) and atrophy (Chiu et al., 2021), although these findings remain controversial (Galougahi et al., 2020; Shor et al., 2021). Given that anosmia is also a common feature of Parkinsonian disease, in which chronic neuroinflammation is thought to play a role, it has been suggested that patients could be at risk of parkinsonism following infection with COVID-19 (Beauchamp et al., 2020). Therefore, it remains important to monitor the olfactory system in COVID-19 survivors given the potential for long-term post-viral Parkinsonism (Beauchamp et al., 2020; Sulzer et al., 2020). With respect to the UKB protocol, the sequence has been shortened by 80 seconds, mainly by increasing the in-plane acceleration factor from R=2 to R=3, keeping the high spatial resolution unchanged.

#### Diffusion-weighted MR Imaging (dMRI)

This sequence measures the ability of water molecules to move within their local environment. The UKB dMRI sequence includes 100 diffusion-encoding directions across 2 b-shells, enabling measurement of the random motion of water molecules to infer information about WM microstructural properties and delineate the gross axonal organisation of the brain. In this multiorgan protocol, we acquired just 3 orthogonal diffusion directions, as commonly done in clinical practice. This allowed us to substantially decrease the acquisition time (from 7:08 to 1:33), while still being able to estimate mean diffusivity (MD), important for example in the assessment of ischaemic injury. Cases of brain ischaemic injury, acute disseminated encephalomyelitis, and encephalitis have been reported in COVID-19 cases (Ellul et al., 2020), while Lu et al, reported significant changes in dMRI-derived measures, including MD, in COVID-19 with respect to controls (Lu et al., 2020).

#### Susceptibility-weighted MR Imaging (swMRI)

This sequence is sensitive to magnetised tissue constituents, including iron, calcium and iron, based on their shifted magnetic susceptibility compared to water (Mittal et al., 2009). Here, we process the magnitude images to calculate T_2_* maps (which reflect compartmentalisation of these constituents) and quantitative susceptibility maps (QSM, which reflect the mean magnetic susceptibility) (Wang et al., 2020). SwMRI is commonly used in clinical settings to study diverse pathologic conditions, such as traumatic brain injury, stroke, and haemorrhages. It can also be used to define vascular malformations, brain tumours, cerebral microbleeds, intracranial calcifications, and iron deposits. Haemorrhagic lesions and extensive and isolated WM microhemorrhages have frequently been reported in COVID-19 (Conklin et al., 2021; Egbert et al., 2020; Ellul et al., 2020; Kremer et al., 2020). The highest iron concentration in the adult brain is found in the basal ganglia and is known to increase with age (Harder et al., 2008). However, focal accumulation of iron is associated with neurodegenerative disorders and has been linked to inflammation (Ward et al., 2014). The only change in this sequence with respect to the one used in the UKB protocol is a small reduction in in-plane spatial resolution (from 0.8×0.8 mm iso to 0.9×0.9 iso), shortening the acquisition time by approximately 30 seconds.

#### Arterial Spin Labelling (ASL)

Brain perfusion has not previously been assessed in UKB, although the recently-initiated UKB post-COVID-19 imaging study has for the first time included a fast, modest ASL protocol. For C-MORE we included a similar multi-slice multi-post label delay (multi-PLD) pseudo-continuous ASL (PCASL) sequence with pre-saturation and two global inversion pulses for background suppression, but here using a 1.4 s labeling duration and a with multi-slice 2D EPI readout (Okell et al., 2013) with 45.2 ms to acquire each slice in ascending order. This sequence is used to quantify tissue perfusion, i.e., cerebral blood flow (CBF), and Arterial Transit Time (ATT) by using the water in arterial blood as an endogenous contrast agent. Given the evidence of vascular damage (haemorrages, strokes, microbleeds, vascular lesions) in some COVID-19 patients (Ellul et al., 2020) in the acute setting, we deemed it to be particularly important to assess brain perfusion in this protocol.

### Dataset: the C-MORE study

This protocol was first used in the C-MORE study. Details of the study are described elsewhere (Raman et al., 2021). Briefly, 58 patients hospitalised for COVID-19 (laboratory-confirmed) between 14th March and 25th May 2020, and 30 uninfected controls (SARS-CoV-2 immunoglobulin negative and asymptomatic), group-matched for age, sex, body mass index (BMI), and risk factors (smoking, diabetes, and hypertension) were included in the study.

The multiorgan MRI scan was carried out 2-3 months from disease onset (median 2.3, IQR 2.06-2.53 months), imaging the brain, lungs, heart, liver and kidneys (3T Prisma, Siemens Healthineers, Erlangen, Germany). The total acquisition time was 70 minutes.

Details on clinical symptoms or signs, vitals and laboratory findings during admission were extracted from electronic medical records, while additional evaluations were performed at the time of scan (see (Raman et al., 2021) for details). The severity of disease during hospital admission was graded as per the WHO ordinal scale for clinical improvement. This study was registered at ClinicalTrials.gov (NCT04510025) and approved in the United Kingdom by the North West Preston Research Ethics Committee (reference 20/NW/0235).

### Analysis pipeline

In this section, we describe the IDPs derived from the automated pipeline. When applicable, we detail pipeline modifications with respect to UKB. The main UKB pipeline is openly available (https://www.fmrib.ox.ac.uk/ukbiobank/) and new IDPs will be included in the next version. Modified or additional scripts and support data for the analyses performed in this study are available at (https://git.fmrib.ox.ac.uk/ludovica/c-more-brain-mri). Examples of pipeline outputs for each modality are shown in Figure 1.

**Figure 1.**
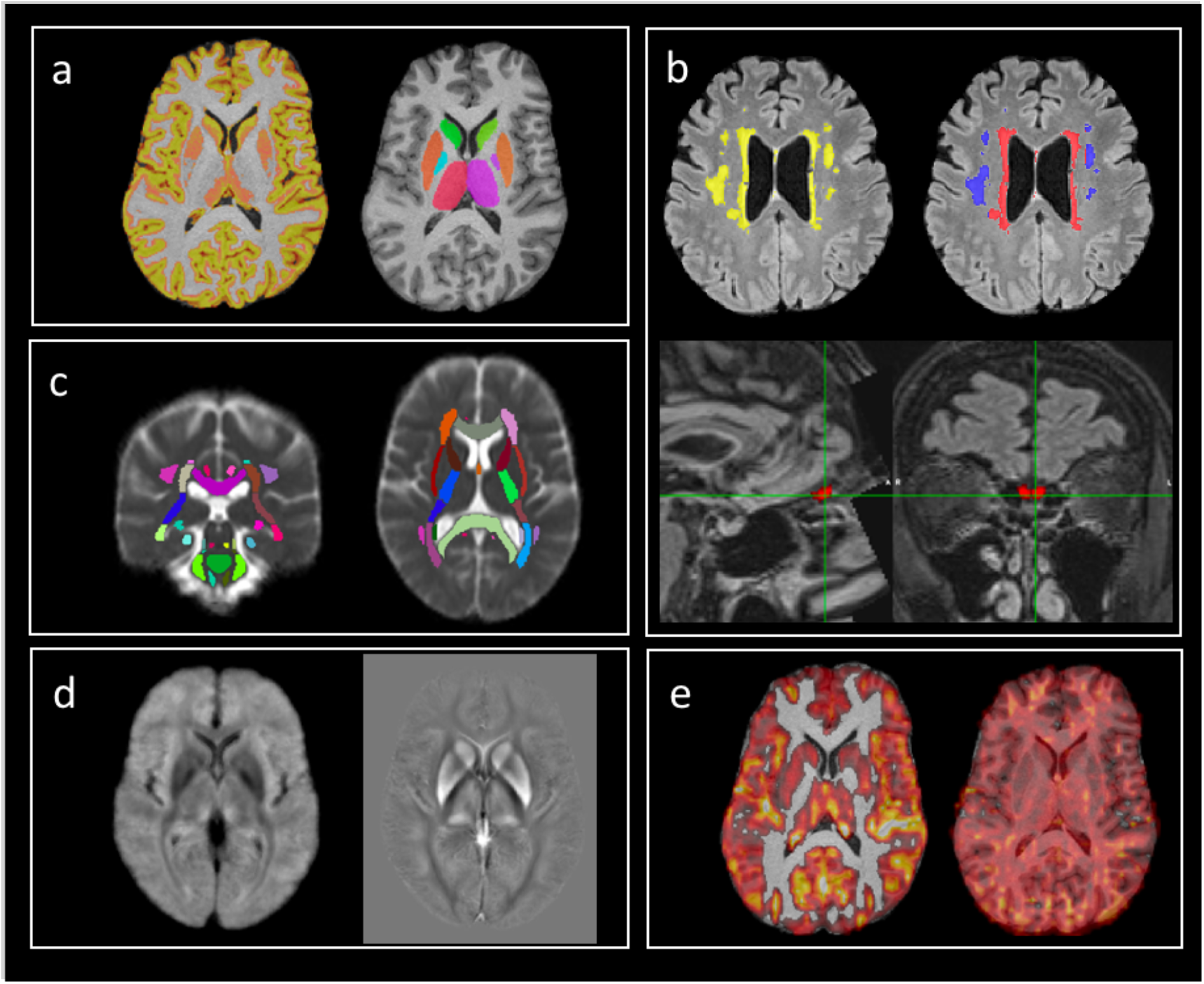
MRI modalities and examples of outputs of the analysis pipeline. a. T_1_: cortical volumes (left - GM partial volume estimate shown as example) and subcortical volumes (right); b. T_2_-FLAIR: white matter hyperintensities (total in yellow, periventricular in red, deep in blue) and olfactory bulb volume and intensity; c. dMRI: mean diffusivity of major white matter tracts (average MD image across subjects shown for display purposes); d. Susceptibility weighted imaging (average images across subjects shown for display purposes): T_2_* (left) and QSM (right); e. ASL: non partial volume corrected cerebral blood flow (left – lower threshold set to 20 ml/100g/min for display purposes) and arterial transit time (right).

Image processing was largely based around tools from FSL (FMRIB Software Library)(Jenkinson et al., 2012). Data were corrected for gradient and EPI distortions and aligned with each other using linear alignment (Jenkinson and Smith, 2001) (between modalities, within-subject). Subjects were then aligned into standard template space (MNI152) using FNIRT nonlinear alignment driven by T_1_-weighted images (Andersson et al., 2019) and aligned to a study-specific template using MMORF nonlinear alignment driven by both the T_1_ and T_2_-FLAIR images (Lange et al., 2020).

#### T_1_ and T_2_-FLAIR

As part of the UKB pipeline, several global and regional volume measures are extracted from the T_1_ scans with SIENAX (Smith et al., 2002). T_1_ images are segmented probabilistically into different tissue types with FAST (Zhang et al., 2001), and from the grey matter partial volume estimates (PVE) (Figure 1a) the average volume (GM density) is calculated within 139 ROIs (defined by the combination of parcellations from several atlases: Harvard-Oxford cortical and subcortical atlases, and Diedrichsen cerebellar atlas, see Supplementary Table S1). The volume of 15 subcortical structures is extracted using FIRST (Patenaude et al., 2011) (Figure 1a).

The T_1_ images are also processed with FreeSurfer using T_2_-FLAIR in conjunction with T_1_ for modelling the cortical surface (Desikan et al., 2006; Fischl et al., 2004). Derived IDPs include subcortical segmentation metrics (Fischl et al., 2002; Iglesias et al., 2015), regional surface area, volume and mean cortical thickness from different parcellations (Desikan-Killiany, Brodmann, Desikan-Killiany-Tourville DKT(Klein and Tourville, 2012), Destrieux), and grey-white intensity contrasts (expressed as the fractional contrast between white and grey matter intensities as sampled either side of the grey-white cortical boundary)(Smith, 2020).

The total volume of WMHs was calculated with BIANCA (Griffanti et al., 2016) using both T_1_ and T_2_-FLAIR images and the UK Biobank training file (available from the UKB pipeline). WMH masks were visually inspected and manually edited if needed, blind to diagnosis (Figure 1b).

In addition to the UKB IDPs, periventricular WMH (PWMH) and deep WMH (DWMH) volumes, defined as being less or more than 10 mm distant from the lateral ventricles, respectively, (DeCarli et al., 2005; Griffanti et al., 2018) (Figure 1b), were extracted as subsets of WMHs.

T_1_ and T_2_-FLAIR images were also used to extract two IDPs to assess potential abnormalities in the olfactory bulb (OB) (Figure 1b). This was achieved in three steps. In the first step, a multimodal, nonlinear template was constructed from T_1_ and T_2_-FLAIR images of 25 UKB participants with a mean age of 63±8 years and 15/25 (60%) men. Average intensity and shape templates were generated with an iterative, multi-resolution approach (Arthofer et al., 2021; Fonov et al., 2011), by minimising both the intensity difference between the template and each subject and the deformation required to warp the template to each subject. The first iteration was initialised with unbiased, affine T_1_ and T_2_-FLAIR templates which were constructed from the same set of subjects and rigidly aligned to the 6th generation nonlinear MNI 152 template (Grabner et al., 2006). Rigid and affine registrations used to align T_1_ and T_2_-FLAIR images within subjects, and T_1_ images between subjects, respectively, were performed with FLIRT (Jenkinson and Smith, 2001). Nonlinear registrations between each subject and the template were performed with MultiModal Registration Framework (MMORF) (Lange et al., 2020), which allowed the simultaneous registration of both modalities, and brain and non-brain tissue. In the second step, the average templates served as a reference space for normalisation and the enhanced SNR enabled clear visualisation of the left and right olfactory bulbs (OB), which were manually segmented. MMORF was used to estimate a deformation field between each individual C-MORE subject’s T_1_ and T_2_-FLAIR images and the template. Finally, each individual’s inverse transformation was used to transform the OB label maps from template space back to each individual’s native space, where the OB volume and OB mean T_2_-FLAIR intensity (normalised with respect to the median T_2_-FLAIR intensity in the white matter) measurements were performed.

#### Diffusion-weighted MR Imaging (dMRI)

Diffusion data were pre-processed with eddy_correct and topup to remove effects of eddy currents, head motion, and susceptibility-induced distortions (Andersson et al., 2003; Jenkinson et al., 2002), and then processed to generate a mean diffusivity (MD) map. The UKB TBSS pipeline was adapted to take into account the availability of MD only. MD images were registered to MNI space combining a linear transformation from the b=0 image to T_1_ with the non-linear registration from T_1_ to MNI. Average MD was then calculated within an average skeleton in standard space for each tract of the JHU atlas (Figure 1c). In addition, the analysis was repeated after excluding voxels where WMHs were present, to assess MD in the normal-appearing WM (NAWM). To achieve this, the linear transformation between T_2_-FLAIR and the b=0 image was calculated and applied to WMH maps. WMHs were then masked out from MD before calculating the mean values in the skeleton’s tracts.

#### Susceptibility-weighted MR Imaging (swMRI)

Both magnitude and phase data acquired from swMRI acquisitions (Figure 1d) were processed to provide quantitative measures reflecting clinically-relevant tissue susceptibility properties. First, as part of the UKB pipeline, magnitude data from two echoes were processed to provide a quantitative mapping of T_2_* signal decay times. Median T_2_* was calculated within each of 14 major subcortical GM structures (previously derived from the T_1_) as IDPs. Second, phase data were processed for QSM following a pipeline being developed for UKB (Wang, 2020). Briefly, phase images from each channel were combined using the MCPC-3D-S approach (Eckstein et al., 2018), the channel-combined phase images from two echoes were unwrapped using a Laplacian-based algorithm (Schofield and Zhu, 2003) and subsequently combined into one phase image via a weighted-average (Wu et al., 2012). This phase image was then filtered using the V-SHARP algorithm (Schweser et al., 2011) to remove background field, and susceptibility maps were generated using the iLSQR algorithm (Li et al., 2015). Voxels within the ventricles were extracted using a ventricle mask derived from the T_2_-FLAIR data (using make_bianca_mask) and subsequently, a mixture modelling algorithm (including one Gaussian and two inverse-Gamma models) (Llera et al., 2016) was applied to the extracted voxels. Susceptibility value of CSF was calculated as the mean value of the main Gaussian distribution and was used as the reference for susceptibility maps. Median susceptibility (CSF-referenced) was calculated in the same 14 subcortical structures as the T_2_* IDPs.

#### Arterial Spin Labelling (ASL)

The ASL data were processed using the BASIL ASL tools in FSL to estimate CBF and ATT (Figure 1e). BASIL analysis involved motion correction, distortion correction (using the field map derived from the blip-up/down diffusion data), brain masking, label-control subtraction, and kinetic model fitting, including modelling of the macrovascular component (Chappell et al., 2008; Chappell et al., 2010) and partial volume correction (PVEc)(Chappell et al., 2011). Absolute CBF quantification was achieved via voxel-wise calibration using the first volume of the ASL data, which was acquired with no ASL preparation or background suppression (the M0 calibration volume). The resulting voxel-wise perfusion images were linearly aligned to the T_1_ structural image using FLIRT with the BBR cost function. Grey matter and white matter PVE maps from FAST were transformed into native ASL space using applywarp with super sampling to integrate the tissue partial volume contributions across the larger ASL native space voxel volumes. A grey matter mask was defined by applying a partial volume threshold of 50% in the native ASL space; this mask was then used to estimate mean grey matter CBF and ATT from both the non-PVEc and PVEc results. Similarly, a white matter mask was defined using a partial volume threshold of 80%; this mask was then used to estimate mean white matter CBF and ATT.

In summary, the current version of this pipeline allows extraction of 1575 IDPs (see complete list in Supplementary Table S1)

### Quality control and neuroradiology reports

Qualitative assessment of all brain MRI images from the C-MORE dataset was undertaken by an expert neuroradiologist (FS), who commented on the presence of white matter hyperintensities, brain atrophy, ischaemic or haemorrhagic abnormalities, and olfactory bulb size and signal.

All data and main outputs of the analysis pipeline were also manually quality-control checked using the same criteria as the UKB imaging study (details in (Alfaro-Almagro et al., 2018)) and an additional set of visual checks for ASL data^1^.

### Exploratory analyses on the C-MORE dataset

As an initial step, we examined the level of agreement between outputs from our automated pipeline against the visual scores provided in the radiology reports. For small vessel disease we compared total WMH volume against visual ratings (none/mild/moderate) using the Kruskal-Wallis test. For brain atrophy we compared total brain volume, total GM and cortical GM (all normalised for head size) between scans classified as normal for age and those reported as showing generalised atrophy using the Mann-Whitney test. Finally, we looked at the volume and intensity on T_2_-FLAIR in the olfactory bulbs.

Group comparisons of IDPs were then undertaken after Gaussianisation (quantile normalisation) of all continuous variables and after regressing out the following confounds: age, sex, BMI, diastolic and systolic blood pressure, smoking and head size. For the FreeSurfer IDPs, the comparisons were based on the Desikan-Killiany-Tourville DKT (Klein and Tourville, 2012) atlas. The conventional level of statistical significance of 5% was used without correction for multiple comparisons. Statistical analyses were performed using SPSS Version 27.0 (IBM, Armonk, NY, USA).

We first explored differences in the IDPs between recovered COVID-19 patients and controls (two sample independent t-test). We then defined as critical those patients who received organ support (any of the following: positive airway pressure ventilation, intubation, dialysis, vasopressor support) and explored differences across severity groups with one-way ANOVA, 3 groups: critical, non-critical, controls (HC).

To look at potential brain changes linked to the loss of smell, we performed a group comparison (ANOVA 3 groups: patients with anosmia, patients without anosmia, controls) on the following IDPs: OB volume and intensity, GM metrics (density, volume, area, cortical thickness) of the amygdala and frontal areas (frontal orbital, frontal medial cortex, frontal pole), brainstem volume.

Follow-up Pearson’s correlations with inflammatory markers (from the blood assay) were performed within the patient group(s) to aid the interpretation of significant group differences.

## RESULTS

Eighty participants completed the brain sequences of the multiorgan scan. After quality control we excluded four participants (2 for excessive motion on T_1_, 1 for presence of an extensive lesion compromising registrations, 1 for pipeline failure at brain extraction level) obtaining usable brain imaging data from 51 patients and 25 controls.

Table 2 summarises the main demographic and disease information from the final 76 participants (95% of the initial sample), together with the number of usable scans per group for each sequence (modality). We also detail the number of radiology reports where a specific comment was made with respect to the amount of small vessel disease, brain atrophy, and olfactory bulbs’ characteristics.

**Table 2.**
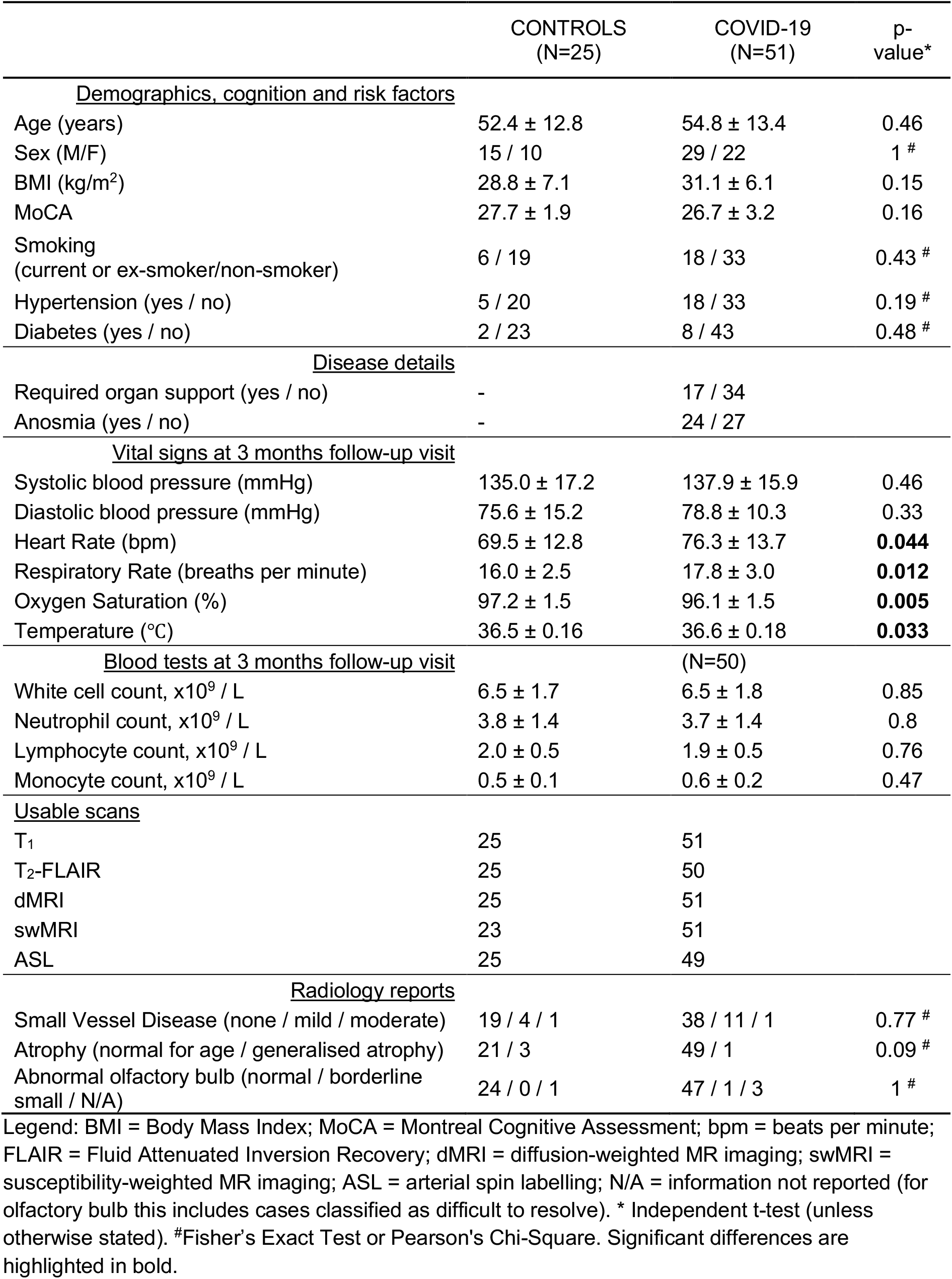
Demographics, vital signs and blood test at follow-up (3 months visit), disease details, usable brain MRI scans, and radiology reports details of patients and control participants in the C-MORE study.

The groups were well matched in terms of demographics and risk factors and showed no significant difference in the patterns of radiological signs. The COVID-19 patients had mildly worse vital signs (higher heart rate and respiratory rate, lower oxygen saturation, and higher body temperature) than controls, although still within the normal range.

### Comparison between IDPs and neuroradiology reports

Figure 2 shows the comparison between IDP values for WMH, total GM volume and OB volume and intensity and the classifications from the radiology reports (independently from diagnosis group). For one participant there was no explicit comment on small vessel disease or atrophy. Most of the scans (78%) were classified as normal. Significant differences in IDPs were observed from the Kruskal-Wallis and Mann-Whitney tests across small vessel disease (SVD) classes and between atrophy classes respectively (Supplementary Table S2), suggesting that IDPs are in line with radiology reports. However, this is only a first validation, which needs to take into account the low number of subjects especially in the “moderate SVD” and “generalised atrophy” classes.

**Figure 2.**
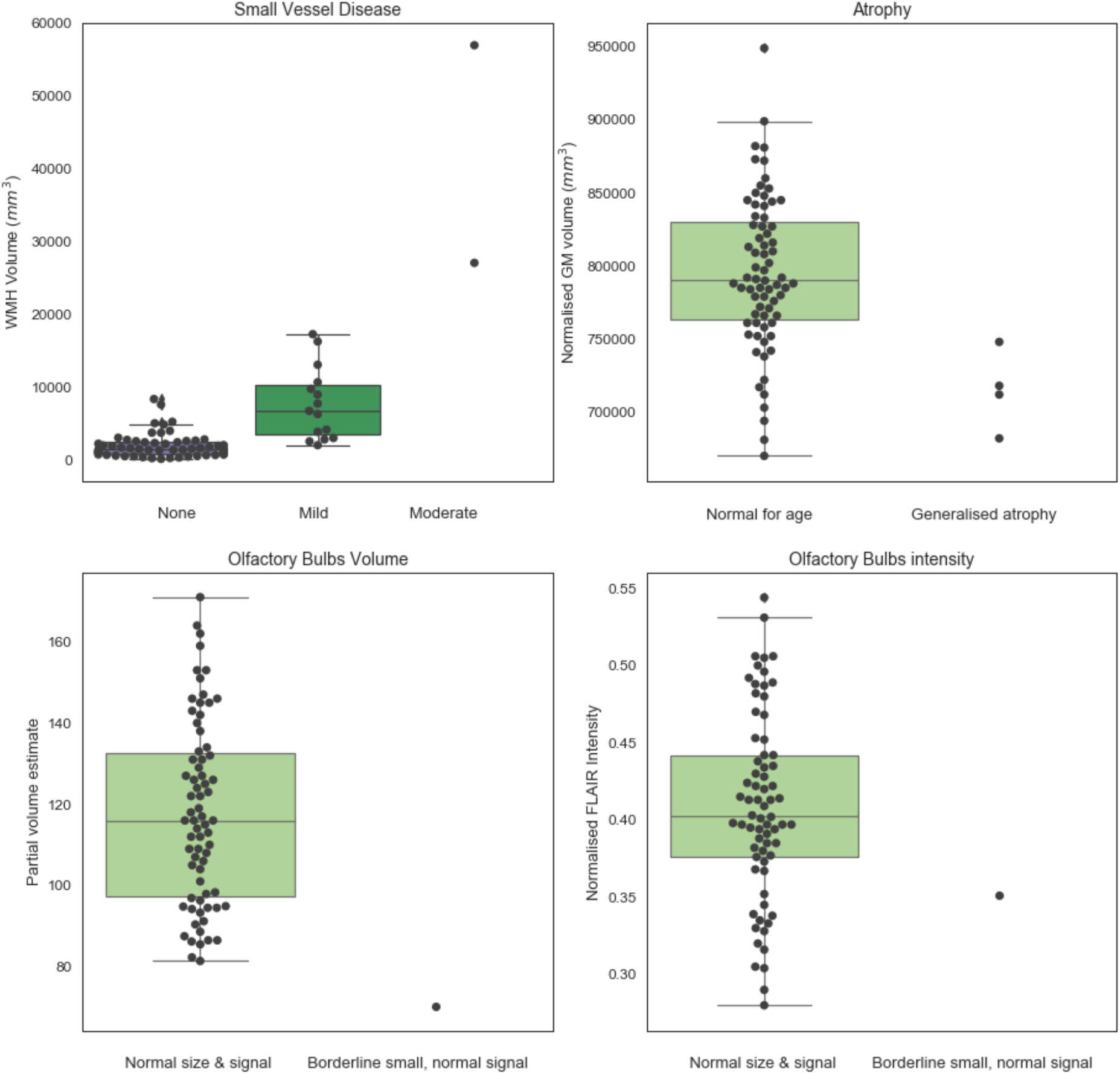
Values of IDPs derived from patients and controls from the automated pipeline against the classification obtained from clinical radiology reports (blind to diagnosis and results of the pipeline). Numerical values are reported in Supplementary Table S2.

Regarding the olfactory bulb, for four participants there was no explicit comment on the size or intensity, or the OB was rated as “difficult to resolve”. The remaining scans were rated as normal in size and signal, with one scan classified as borderline small, but with normal signal, in line with the IDPs relative to OB volume (lower than all scans rated as normal) and T_2_-FLAIR intensity (value within the 20^th^ percentile of the scans rated as normal) (Figure 2, bottom row).

### Results of the cohort comparisons based on disease group

Figure 3 summarises the key results of our exploratory analyses. Supplementary Tables S3 and S4 detail all the significant group differences that we observed between previously hospitalised COVID-19 patients and controls (Table S3, with anatomical areas shown in Supplementary Figure S1) and across severity groups (Table S4, with anatomical areas shown in Supplementary Figure S2): patients who required organ support (critical), patients who did not require organ support (non-critical), controls (HC).

**Figure 3.**
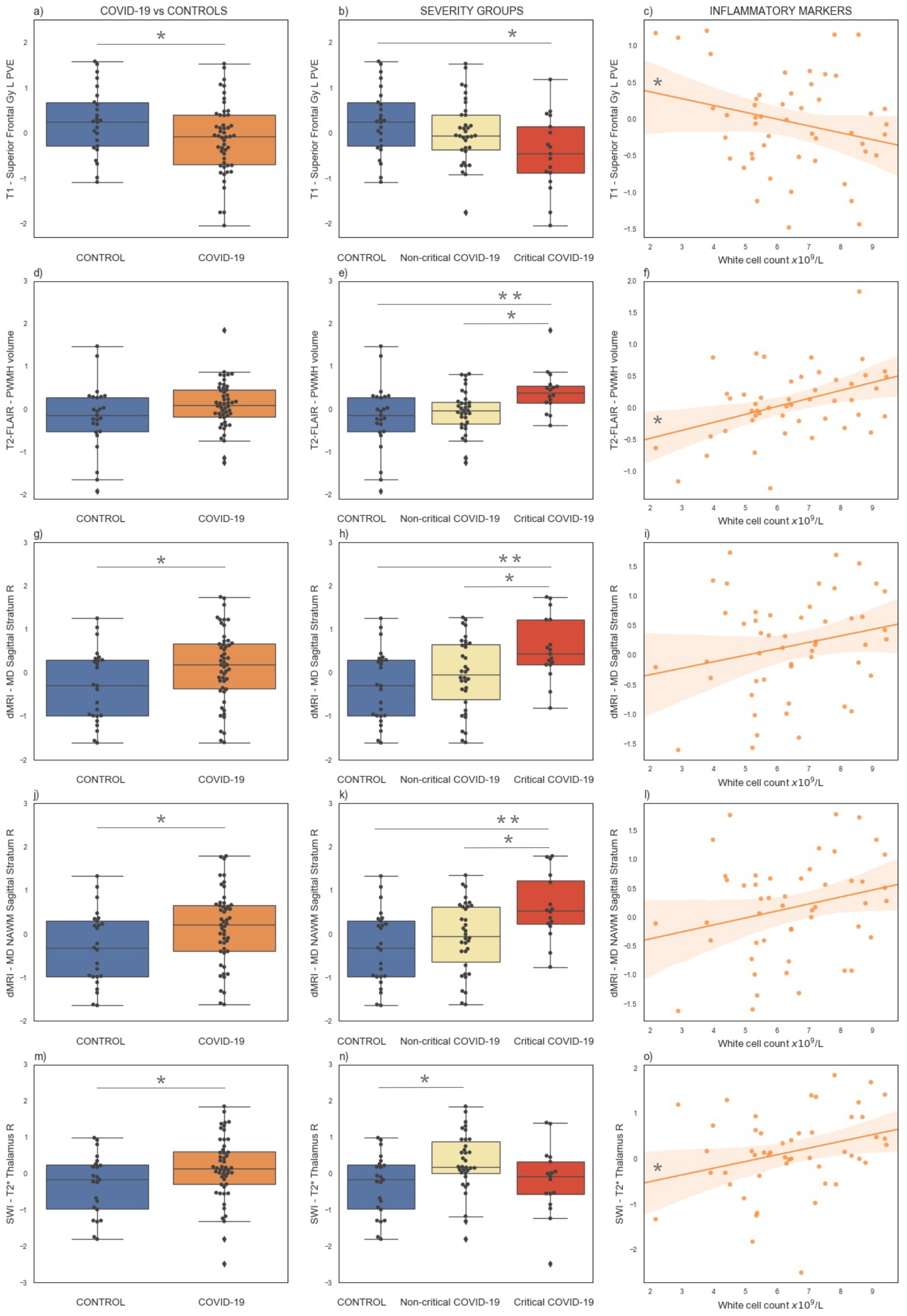
Key results of the exploratory analyses on multimodal brain MRI-derived IDPs from the C-MORE study. The first column (Panels a,d,g,j,m) shows the comparison between COVID-19 patients and controls. The middle column (Panels b,e,h,k,n) shows the comparison across severity groups. The right column (Panels c,f,i,l,o) shows the relationship between IDPs and inflammatory markers (white cell count) in COVID-19 survivors. All IDPs were Gaussianised and deconfounded. * p<0.05, **p<0.01. See supplementary material for corresponding raw values, details of all significant results and anatomical location of the brain areas involved.

#### T_1_ and T_2_-FLAIR

The main differences in the grey matter between COVID-19 patients and controls were observed in the frontal lobe. COVID-19 patients had lower GM density in the left superior frontal gyrus (Figure 3a) and a lower volume and mean cortical thickness in the caudal portion of the left middle frontal gyrus. The GM density and volume of the left superior frontal gyrus, the GM density of the right inferior frontal gyrus (pars triangularis), the cortical thickness of the left lateral orbitofrontal gyrus and the cortical thickness of the left pars orbitalis of the inferior frontal gyrus were significantly different across severity groups, with lowest GM volume in the patients who received organ support (Figure 3b) (details in Supplementary Tables S3-S4). Within the COVID-19 patients, the GM density of the left superior frontal gyrus was negatively correlated with systemic markers of inflammation (white cell count: r=−0.34, p=0.015; neutrophils count: r=−0.28, p=0.049; lymphocytes count: r=−0.33, p=0.019; monocytes count: r=−0.31, p=0.03) (Figure 3c) and the cortical thickness of the left lateral orbitofrontal cortex was negatively correlated with lymphocytes count (r=−0.32, p=0.02). When considering critical COVID-19 patients only, the GM density of the left superior frontal gyrus was negatively correlated with lymphocytes count (r=−0.50, p=0.04), while the volume was positively correlated with monocytes count (r=0.59, p=0.01). No significant correlations were found in the non-critical group.

In addition, COVID-19 patients had decreased GM metrics (GM density, volume, thickness, or area) with respect to controls in the following areas: left hippocampus (lower volume), left superior division of the lateral occipital cortex (lower GM density), transverse temporal gyrus (lower left volume), right middle temporal gyrus (lower thickness, lowest in the non-critical group), right superior temporal gyrus (lower thickness), right inferior parietal (lower thickness), right supramarginal gyrus (lower thickness, lowest in the non-critical group), right isthmus of the cingulate gyrus (lower area and volume, lowest in the non-critical group), right cuneus (lower thickness) (details in Supplementary Tables S3-S4). No significant correlations with inflammatory markers were found in these areas within the patient group or severity sub-groups.

The only increase in GM metrics in COVID-19 patients with respect to controls was found in the area of the right transverse temporal gyrus, with the highest values in the non-critical group (not correlated with inflammatory markers).

Critical COVID-19 patients had an increased burden of total WMH, in particular periventricular WMH (Figure 3e). WMH volumes were positively correlated with inflammatory markers in the COVID-19 group (Total WMH with white cell count: r=0.36, p=0.012; Total WMH with lymphocytes count: r=0.39, p=0.006. PWMH with white cell count: r=0.42, p=0.003; neutrophils count: r=0.32, p=0.025; lymphocytes count: r=0.42, p=0.003; monocytes count: r=0.30, p=0.036) (Figure 3f). When splitting the COVID group according to critical status, the correlation between WMH and some inflammatory markers remained significant only within the non-critical group (Total WMH - lymphocytes count: r=0.45, p=0.009; PWMH - lymphocytes count: r=0.49, p=0.004; PWMH - white cell count: r=0.39, p=0.024).

#### Diffusion-weighted MR Imaging (dMRI)

As previously shown by us, COVID-19 patients had increased mean diffusivity in the left posterior thalamic radiation and right sagittal stratum compared to controls (Figure 3g). When comparing severity groups, MD in the right sagittal stratum was higher in patients who received organ support with respect to both non-critical patients and controls (Figure 3h). When looking at NAWM only, the results remained significant in the sagittal stratum (Figure 3j,k) and significantly higher MD was observed in the right posterior thalamic radiation in critical patients with respect to controls. No significant correlation was found with inflammatory markers, although we observed a trend of increased MD corresponding to higher inflammation (Figure 3 i,l).

#### Susceptibility-weighted MR Imaging (swMRI)

COVID-19 patients had a higher T_2_* signal in the thalamus with respect to controls (left p=0.052; right p=0.037) (Figure 3m), lower T_2_* in the left hippocampus and higher QSM in the right hippocampus. The comparison across severity groups revealed that the T_2_* signal in the right thalamus was higher in patients who did not receive organ support with respect to controls (Figure 3n). Within the COVID-19 patients, the T_2_* signal in the right thalamus was positively correlated with inflammation markers (white cell count: r=0.30, p=0.036; neutrophils count: r=0.38, p=0.007) (Figure 3o) and remained significant when considering non-critical COVID-19 patients only (white cell count: r=0.46, p=0.007; neutrophils count: r=0.51, p=0.003).

#### Arterial Spin Labelling (ASL)

No significant group differences were observed in CBF or ATT IDPs.

### Results of the cohort comparisons based on anosmia

When looking at the olfactory system (olfactory bulbs and key GM areas) no significant differences were found across anosmia groups on the selected IDPs.

## DISCUSSION

In this work we describe how we adapted the UKB brain MRI protocol and processing pipeline to multi-organ imaging of COVID-19, with initial direct application to the C-MORE study.

We developed a 17-minute MRI protocol to study the effect of COVID-19 on the brain. By modifying the UK Biobank protocol, we were able to exploit its technical advances to generate high quality data in a reduced time, while tailoring it to the specific purpose of assessing the effects of COVID-19 (e.g. introducing ASL to assess brain perfusion). While some studies used multimodal MRI to look at changes after COVID-19 (Lu et al., 2020), to the best of our knowledge this is the most comprehensive brain imaging protocol adopted so far, including five MRI modalities. This will enable the monitoring of a wide range of potential medium and long-term effects of SARS-CoV-2 virus on the brain. This is also in line with the approach adopted by emerging consortia (Edlow et al., 2021) which aim to develop neuroimaging common data elements that are capable of capturing the broad spectrum of brain imaging findings reported to date in adult patients with COVID-19 and that are feasible to implement in hospitals around the world, as already demonstrated in the UK-wide C-MORE study. In addition, the close similarity of our protocol to the UK Biobank protocol will enable utilisation of that dataset (e.g., for normative co-modelling). We obtained usable data from 95% of participants, suggesting that the protocol is well tolerated and can be used as part of a multiorgan MRI assessment.

The analysis pipeline currently generates 1575 IDPs, including some specifically designed to look at potential effects of COVID-19. In particular, we were able to segment the olfactory bulbs, thanks to improvements in non-brain alignment and template construction, and we generated 8 IDPs from ASL. Many of these IDPs are also suitable for evaluating the feature-based common data elements proposed by GCS-NeuroCOVID for reporting MRI in COVID-19 (Edlow et al., 2021).

For brain characteristics that are commonly reported by neuroradiologists, we compared our IDPs with clinical reports and found reasonable agreement, particularly for small vessel disease and atrophy. This suggests that the pipeline can be used to automatically extract meaningful information from larger datasets.

The first application of this protocol and pipeline was in the C-MORE study. Our exploratory analyses on the IDPs revealed some interesting albeit preliminary group differences.

In the grey matter, we observed localised reduced GM metrics in COVID-19 patients. In the frontal lobe, values were significantly lower in critical patients and often negatively correlated with inflammation markers. Reductions observed in non-frontal areas were instead more prominent in non-critical patients and not correlated with inflammatory markers, suggesting a different pattern of the impact of COVID-19 on the GM. The only increase in GM in COVID-19 patients was observed in the area of the right transverse temporal gyrus, which was more prominent in non-critical COVID-19 patients. Our results are partially in contrast with the findings of Lu and colleagues (Lu et al., 2020), who only reported increases in GM volumes in COVID-19 patients with respect to controls. In addition, observed differences in GM characteristics were in different brain locations to the ones we observed. While the interpretation of these differences in GM volume remains unclear (Goldberg et al., 2021), the negative association of white cell count with grey matter volume suggests that a dysregulated inflammatory response in COVID-19 patients may be linked to changes in grey matter volume. Further investigation is needed to better characterise the cause and extent of this effect.

We specifically looked at the olfactory system (olfactory bulbs and key GM areas) and did not find differences between COVID-19 patients who experienced anosmia during their acute illness versus those who did not and controls. This in line with studies reporting normal OB in anosmic patients (Galougahi et al., 2020; Shor et al., 2021), and with Lu and colleagues (Lu et al., 2020), who did not find significant group differences in patients with and without loss of smell. However, it must be noted that only two patients experienced smell loss in the cohort studied by Lu et al., (Lu et al., 2020) and among the 24 patients with anosmia in our study, the majority of them (N=18) were in the non-critical group. Of importance our analysis was restricted to data collected on anosmia during admission. One possible explanation for the lack of differences in imaging derived metrics from the olfactory system is that the symptoms may have resolved by the time of the scan. Future studies on larger samples might be better able to detect brain changes associated with anosmia or further confirm the lack of a radiological signature of this specific COVID-19 symptom.

In the present study, we also observed some macroscopic as well as microstructural differences in the white matter. COVID-19 patients, particularly critical ones, showed an increased amount of WMHs, especially periventricular WMH. Of interest, a similar increase in WMHs has also been reported by others (Varatharaj et al., 2020) and thought to reflect a higher burden of microvascular damage. While this is biologically plausible, the cross-sectional analysis used in this study limits our ability to make causal inferences.

The increased MD in the sagittal stratum and posterior thalamic radiation (also evident when evaluating normal appearing WM), suggests a disruption of healthy white matter. In fact, MD is typically seen to increase in aging and many diseases. Lu and colleagues also reported some white matter disruption in different tracts of the brain. However, in contrast to us, MD was seen to decrease rather than increase (Lu et al., 2020). While the precise reasons for these differences are unclear, some possible explanations include differences in the dMRI protocol (Lu and colleagues used a higher number of diffusion directions) and confounding variables used in the analyses (in addition to age, sex and head size we also used BMI, blood pressure and smoking). Of interest, longitudinal studies examining the trajectory of changes in these measures could provide the much needed clarity required in interpretation of these findings.

The differences we found in frontal GM, WMHs and MD were stronger in patients who received organ support, suggesting a relationship between disease severity and imaging biomarkers of neurological health. This is in line with a recent cohort study using electronic health records (Taquet et al., 2021) showing a clear effect of COVID-19 severity on subsequent neurological diagnosis. Given the association of brain atrophy, vascular pathology and WM microstructural damage on MRI with neurodegenerative diseases such as dementia, it would be prudent to monitor the long-term effects of such MRI abnormalities on cognitive performance of patients (Miners et al., 2020).

Another important finding in this study is the association of some brain IDPs with inflammatory markers. A recent UK-wide follow up study of previously hospitalised COVID-19 patients (N= 1,077) have identified a subgroup of patients with cognitive impairment, increased markers of inflammation and high symptom burden (PHOSP-COVID Collaborative Group et al., 2021). The correlation of brain IDPs and white cell count (a marker of inflammation) in our study highlights the considerable value that follow-up brain imaging will add to our understanding the mechanisms underlying cognitive impairment in COVID-19 patients.

Neuropsychological and brain changes (including atrophy and WMHs) are known to occur in in patients admitted to ICU and survivors of critical illness (Jackson et al., 2014; Suchyta et al., 2010). Since we also observed brain changes in non-critical patients, the organ support received by critical patients is unlikely to be the cause of all the findings. However, as our controls were not hospitalised, we cannot, with certainty, attribute the observed differences to COVID-19. It is also worth noting that the type of organ support that the 17 C-MORE COVID-19 patients received was quite heterogeneous (positive airway pressure ventilation, intubation, dialysis, vasopressor support, medical therapy), therefore further studies with larger samples may be needed to potentially disentangle effects related to specific interventions. This could potentially include data from non-COVID-19 patients who received organ support as an additional control group.

From susceptibility-weighted scans we observed a significant difference in T_2_* in the thalamus bilaterally, which was positively correlated with white cell count. Pathological alterations in other deep structures are usually observed as reductions in T_2_*, for example, due to higher iron accumulation (Ward et al., 2014). However, this opposite trend between the thalamus and other subcortical structures has been previously observed in UK Biobank data (see (Miller et al., 2016) Figure 8): T_2_* in subcortical structures was found to be negatively correlated with age, while thalamus T_2_* showed a positive correlation with age. The fact that no difference was observed in QSM between COVID-19 patients and controls suggests that this could be a difference in tissue compartmentalisation. Interestingly, T_2_* in the right thalamus was higher in non-critical patients. Potential explanations are that more critical COVID-19 involved other brain changes and not the thalamus, or that the treatment (e.g. steroids) received by critical patients contributed to normalising T_2_* signal in this area. We also observed decreased T_2_* and increased QSM in the hippocampus in COVID-19 patients, which could be due to higher iron accumulation related to the virus infection. However, measurements in the hippocampus are likely to be affected by partial volume effect, due to the small size of the area and relatively thick slices. This is also an area that is prone to MRI signal dropout, so we cannot exclude this potential confound.

This exploratory study has several limitations. The sample size is relatively small and, due to the comparisons performed on a wide range of IDPs, the results do not survive correction for multiple comparison. Anosmia was not assessed at the time of scan but rather at admission. Group differences in brain IDPs between COVID subgroups could be related to the therapy used while in hospital and not specific to COVID-19 per se.

In conclusion, we developed a brain MRI protocol and analysis pipeline that enable efficient yet comprehensive assessment of brain changes in COVID-19 patients, and could be used both as part of a multiorgan imaging study as well as standalone. The observed changes in the C-MORE study have provided early insights into the potential repercussions following infection. The C-MORE study included a limited number of previously hospitalised patients and focused on assessing medium-term damage, but has paved the way for more extensive and longitudinal studies that can take advantage of the protocol and analysis developed as part of this study. The follow-up phase of the C-MORE study is currently ongoing and aims to monitor the longitudinal trajectory of multiorgan health beyond the subacute phase. This multiorgan imaging protocol is also now being used in the ‘PHOSP-COVID Post-hospitalisation COVID-19’ study, a large UK-wide national consortium (N=616), and in the CONVALESCENCE study, a study of patients with Long COVID (N=800). This will allow for testing the reproducibility of our preliminary results and will augment our understanding of the medium to long term neurological sequele of COVID-19.

Finally, UK Biobank has just begun imaging 3,000 volunteers from the 45,000 subjects that were already scanned before the pandemic. Half of these will have been invited for scanning because tests show that they have been exposed to the virus and became infected (while the other half will be a control group). This pre-vs post-COVID-19 imaging data will be a large and rich resource for studying the effects of this disease in the brain and other parts of the body, and the preprocessing and analyses of these data will benefit from the work described above.

## Data Availability

Individual de-identified participant data will be made available when the trial is complete, or upon request directed to Dr Raman; after approval of a proposal, data can be shared through a secure online platform. The main UK Biobank brain MRI analysis pipeline is openly available (https://www.fmrib.ox.ac.uk/ukbiobank/) and code for new IDPs will be included in the next version. Modified or additional scripts and support data for the analyses performed in this study are available at (https://git.fmrib.ox.ac.uk/ludovica/c-more-brain-mri).

## Declaration of interests

Dr. Raman reports grants from NIHR Oxford Biomedical Research Centre, grants from United Kingdom Research Innovation Award, and Oxford British Heart Foundation Centre for Research Excellence during the conduct of the study. Dr. Cassar reports grants from NIHR Oxford Biomedical Research Centre, during the conduct of the study. Dr. Tunnicliffe reports grants from NIHR Oxford Biomedical Research Centre, during the conduct of the study; shareholding in Perspectum, outside the submitted work. Dr. Okell reports grants from Wellcome Trust/Royal Society, during the conduct of the study; personal fees from SBGNeuro, personal fees from Oxford University Press, personal fees from Siemens Healthineers, outside the submitted work; In addition, outside the submitted work, Dr. Okell has a patent Combined angiography and perfusion using radial imaging and arterial spin labelling pending, a patent (with Dr. Jezzard) Off-resonance Correction for Pseudo-continuous Arterial Spin Labelling pending, a patent Estimation of blood flow rates issued, a patent (with Dr. Chappell) Fast analysis method for non-invasive imaging of blood flow using vessel-encoded arterial spin labelling with royalties paid by Siemens Healthineers, and a patent (with Dr. Jezzard and Dr Chappell) Quantification of blood volume flow rates from dynamic angiography data with royalties paid by Siemens Healthineers. Dr. Chappell reports personal fees from Oxford University Press and Springer-Nature, outside submitted work. Dr. Neubauer reports grants from Oxford NIHR Biomedical Research Centre, grants from UKRI, during the conduct of the study; personal fees, and others from Perspectum Diagnostics, outside the submitted work. Dr. Jezzard reports grants from Oxford NIHR Biomedical Research Centre and salary support from the Dunhill Medical Trust. Dr. Griffanti, Dr. Alfaro-Almagro, Dr. Chappell, Dr. Andersson, and Dr. Smith receive royalties from licensing of FSL to non-academic, commercial entities. All other authors have no competing interests to declare.

## Author contributions

*Ludovica Griffanti* - Conceptualization, Methodology, Software, Investigation, Data Curation, Formal analysis, Visualization, Writing - Original Draft

*Betty Raman* - Conceptualization, Investigation, Data Curation, Project administration, Funding Acquisition, Writing - Review & Editing

*Nicola Filippini* - Investigation, Data Curation, Formal analysis, Visualization, Writing - Review & Editing

*Fidel Alfaro-Almagro* - Software, Data Curation, Formal analysis, Writing - Review & Editing

*Mark Philip Cassar* - Conceptualization, Investigation, Data Curation, Project administration, Writing - Review & Editing

*Fintan Sheerin* - Methodology, Investigation, Writing - Review & Editing

*Thomas Okell* - Software, Investigation, Writing - Review & Editing

*Flora A Kennedy McConnell* - Software, Investigation, Writing - Review & Editing

*Michael A Chappell* - Software, Investigation, Writing - Review & Editing

*Chaoyue Wang* - Software, Investigation, Writing - Review & Editing

*Christoph Arthofer* - Software, Investigation, Writing - Review & Editing

*Frederik J Lange* - Software, Investigation, Writing - Review & Editing

*Jesper Andersson* - Methodology, Software, Writing - Review & Editing

*Clare E Mackay* - Resources, Funding Acquisition, Writing - Review & Editing

*Elizabeth Tunnicliffe* - Methodology, Software, Writing - Review & Editing

*Matthew Rowland* - Methodology, Investigation, Writing - Review & Editing

*Stefan Neubauer* - Conceptualization, Methodology, Supervision, Funding Acquisition, Writing - Review & Editing

*Karla Miller* - Conceptualization, Methodology, Investigation, Writing - Review & Editing

*Peter Jezzard* - Conceptualization, Methodology, Writing - Review & Editing

*Stephen Smith* - Conceptualization, Methodology, Investigation, Resources, Supervision, Funding Acquisition, Writing - Review & Editing

## Acknowledgments

We thank our patients and their families who have sought to help others understand the effects of COVID-19. We are grateful to the University of Oxford and Oxford University Hospital Trust for their support of this study. We would like to acknowledge OCMR staff, Ms Rebecca Mills, Ms Polly Whitworth, Ms Joana Leal Pelado, Ms Claudia Nunes, Ms Harriet Nixon, Ms Injung Jang, Ms Maryam Khan, Ms Gillian Roberts, Mr Mike MacDonald, Ms Jasmine Taylor, Dr Ahmad Moolla, Ms Sarah White, Dr Ines Abdesselam, for their help with this work. We thank Hanan Lamlum for her help in application of ethics. We acknowledge the support of Siemens in providing a licence for the use of ASL. The diffusion sequence used in our protocol was kindly provided by the CMRR, University of Minnesota, USA.

## Funding sources

NIHR Oxford and Oxford Health Biomedical Research Centres, British Heart Foundation Centre for Research Excellence, UKRI, Wellcome Trust, British Heart Foundation. The authors’ work was supported by NIHR Oxford and Oxford Health Biomedical Research Centre, Oxford British Heart Foundation (BHF) Centre of Research Excellence (RE/18/3/34214), United Kingdom Research Innovation and Wellcome Trust. This project is part of a tier 3 study (C-MORE) within the collaborative research programme entitled PHOSP-COVID Post-hospitalisation COVID-19 study: a national consortium to understand and improve long-term health outcomes. Funded by the Medical Research Council and Department of Health and Social Care/ National Institute for Health Research Grant (MR/V027859/1) ISRCTN number 10980107. This work also arises from one of the national “COVID-19 Cardiovascular Disease Flagship Projects” designated by the NIHR-BHF Cardiovascular Partnership. PJ thanks the Dunhill Medical Trust for support. MAC and FKM acknowledge support from EPSRC (EP/P012361/1). TWO was supported by a Sir Henry Dale Fellowship jointly funded by the Wellcome Trust and the Royal Society (220204/Z/20/Z). The Wellcome Centre for Integrative Neuroimaging is supported by core funding from the Wellcome Trust (203139/Z/16/Z). The authors gratefully acknowledge funding from the Wellcome Trust Collaborative Award (215573/Z/19/Z). KLM acknowledges further support from the Wellcome Trust Senior Research Fellowship (202788/Z/16/Z).

## Footnotes

1 https://git.fmrib.ox.ac.uk/ludovica/c-more-brain-mri/-/tree/master/ASL_pipeline

## Notes

### Clinical Trial

This study was registered at ClinicalTrials.gov, NCT04510025

### Author Declarations

This study was approved in the United Kingdom by the North West Preston Research Ethics Committee (reference 20/NW/0235).

