## Supplementary for "Adapting the UK Biobank brain imaging protocol and analysis pipeline for the C-MORE multi-organ study of COVID-19 survivors": GriffantiRaman_Supplem2 C-MORE brain.pdf

**Supplementary Table S1**: See file Supplem_table1_IDPs_list.xlsx

**Supplementary Table S2**: Values of IDPs derived from the automated pipeline against the classification obtained from radiology reports (blind from diagnosis and results of the pipeline).

| Imaging Derived Phenotypes (IDPs) | Classification from radiology report | | | |
|---|---|---|---|---|
| Small Vessel Disease | None (N=57) | Mild (N=15) | Moderate (N=2) | p-value (Kruskal-Wallis test) |
| WMH total volume | 1497 (766 - 2529) | 6758 (3044 - 10715) | 27066 - 56980 | <0.001 |
| Atrophy | Normal for age (N=71) | Generalised atrophy (N=4) | | p-value (Mann-Whitney test) |
| Total brain volume (normalised for head size) | 1564742.10 (1503956.50 - 1604987.20) | 1421873.85 (1396672.03 - 1445727.93) | | 0.003 |
| Total GM volume (normalised for head size) | 789955.41 (761463.80 - 833483.23) | 714944.74 (689539.57 - 740432.56) | | 0.002 |
| Cortical GM volume (normalised for head size) | 641676.86 (610883.32 - 669832.62) | 582385.24 (525636.60 - 607881.44) | | 0.007 |
| Olfactory bulbs | Normal (N=71) | Borderline small, normal signal (N=1) | | |
| Partial volume estimate | 116.10 (96.86 - 133.15) | 70.40 (<1st percentile) | | |
| $T_2$-FLAIR intensity (normalised for WM FLAIR intensity) | (N=70) 0.40 (0.37 - 0.44) | 0.35 (20th percentile) | | |

Legend: WMH = white matter hyperintensities; GM = grey matter; FLAIR = Fluid-attenuated inversion recovery; WM = white matter.

**Supplementary Table S3.** Results of exploratory group comparisons (COVID vs CONTROLS) on the IDPs generated by the pipeline.

| | CONTROLS | | | | COVID-19 | | | | p-value |
|---|---|---|---|---|---|---|---|---|---|
| | N | Median | 25 | 75 | N | Median | 25 | 75 | |
| T1_GM_parcellation_L_Sup_Front_Gyr_vol | 25 | 11492.7 | 10274.1 | 12940.1 | 51 | 10536.0 | 9639.6 | 11545.6 | 0.042 |
| T1_GM_parcellation_L_Lateral_Occ_Sup_vol | 25 | 16782.2 | 15939.5 | 19658.5 | 51 | 15831.4 | 14393.9 | 18151.2 | 0.033 |
| T1_GM_parcellation_L_Hippocampus_vol | 25 | 4207.9 | 3823.2 | 4345.5 | 51 | 3868.4 | 3591.2 | 4158.9 | 0.018 |
| aparc-DKTatlas_lh_thickness_caudalmiddlefrontal | 25 | 2.865 | 2.7985 | 2.949 | 51 | 2.794 | 2.717 | 2.87 | 0.023 |
| aparc-DKTatlas_lh_volume_caudalmiddlefrontal | 25 | 7390 | 6811.5 | 7965.5 | 51 | 6788 | 6137 | 7267 | 0.026 |
| aparc-DKTatlas_lh_volume_transversetemporal | 25 | 1300 | 1208.5 | 1379.5 | 51 | 1172 | 1037 | 1302 | 0.043 |
| aparc-DKTatlas_rh_area_transversetemporal | 25 | 313 | 295.5 | 333.5 | 51 | 324 | 297 | 351 | 0.011 |
| aparc-DKTatlas_rh_thickness_middletemporal | 25 | 3.025 | 2.952 | 3.1075 | 51 | 2.925 | 2.803 | 3.048 | 0.014 |
| aparc-DKTatlas_rh_thickness_superiortemporal | 25 | 3.145 | 3.0245 | 3.2175 | 51 | 2.976 | 2.894 | 3.149 | 0.046 |
| aparc-DKTatlas_rh_thickness_inferiorparietal | 25 | 2.769 | 2.6525 | 2.8275 | 51 | 2.702 | 2.59 | 2.759 | 0.017 |
| aparc-DKTatlas_rh_thickness_supramarginal | 25 | 2.921 | 2.782 | 2.9745 | 51 | 2.796 | 2.678 | 2.884 | 0.006 |
| aparc-DKTatlas_rh_area_isthmuscingulate | 25 | 976 | 865 | 1050.5 | 51 | 889 | 811 | 982 | 0.017 |
| aparc-DKTatlas_rh_volume_isthmuscingulate | 25 | 2685 | 2550 | 2947 | 51 | 2462 | 2249 | 2723 | 0.004 |
| aparc-DKTatlas_rh_thickness_cuneus | 25 | 2.026 | 1.979 | 2.1065 | 51 | 1.951 | 1.888 | 2.049 | 0.049 |
| dMRI_TBSS_MD_Posterior_thalamic_radiation_L | 25 | 811.0 | 791.5 | 830.5 | 51 | 831.0 | 814.0 | 853.0 | 0.047 |
| dMRI_TBSS_MD_Sagittal_stratum_R | 25 | 813.0 | 787.5 | 829.0 | 51 | 840.0 | 799.0 | 866.0 | 0.026 |
| dMRI_TBSS_MD_NAMW_Posterior_thalamic_radiation_L | 25 | 805.0 | 785.0 | 823.5 | 50 | 826.0 | 813.3 | 846.3 | 0.05 |
| dMRI_TBSS_MD_NAMW_Sagittal_stratum_R | 25 | 813.0 | 786.5 | 829.0 | 50 | 837.0 | 798.5 | 858.5 | 0.027 |
| SWI_T2star_r_thalamus | 23 | 42.4 | 40.0 | 45.2 | 51 | 43.9 | 41.7 | 45.9 | 0.037 |
| SWI_T2star_l_hippocampus | 23 | 45.4 | 43.8 | 47.7 | 51 | 43.5 | 40.8 | 45.9 | 0.041 |
| SWI_QSM_r_hippocampus_refCSF | 23 | -1.4 | -4.7 | 2.9 | 51 | 3.4 | -1.7 | 6.0 | 0.048 |

The table reports results for the IDPs which showed a significant group difference. Median and interquartile range of raw data are presented in the table for ease of interpretation. P-values were derived from independent Student's t-test comparison of gaussianised deconfounded data. Supplementary figure S1 shows the anatomical location of the brain areas involved.

**Supplementary Table S4.** Results of exploratory group comparisons on the IDPs generated by the pipeline across severity groups: COVID patients who received organ support (C19+), COVID patients who did not receive organ support (C19-) and CONTROLS.

| | CONTROLS (HC) | | | | Non-critical COVID-19 (C19-) | | | | Critical COVID-19 (C19+) | | | | F | p-value | Post-hoc multiple comparisons (p-values bonferroni corrected) |
|---|---|---|---|---|---|---|---|---|---|---|---|---|---|---|---|
| | N | Median | p25 | p75 | N | Median | p25 | p75 | N | Median | p25 | p75 | | | |
| T1_GM_parcellation_L_Sup_Front_Gyr_vol | 25 | 11492.7 | 10274.1 | 12940.1 | 34 | 10719.5 | 10121.5 | 11732.4 | 17 | 9753.5 | 7990.5 | 10897.6 | 4.53 | 0.014 | C19+ < HC p=0.011 |
| T1_GM_parcellation_R_Inf_Front_Gyr_pars_triangularis_vol | 25 | 2227.8 | 1884.1 | 2539.8 | 34 | 2383.5 | 2032.5 | 2671.8 | 17 | 1841.4 | 1737.9 | 2284.3 | 4.056 | 0.021 | C19+ < C19- p=0.018 |
| aparc-DKTatlas_lh_thickness_lateralorbitofrontal | 25 | 2.8 | 2.8 | 2.9 | 34 | 2.8 | 2.7 | 2.9 | 17 | 2.7 | 2.7 | 2.8 | 4.616 | 0.013 | C19+ < C19- p=0.01 |
| aparc-DKTatlas_lh_thickness_parsorbitalis | 25 | 2.9 | 2.7 | 2.9 | 34 | 2.9 | 2.8 | 3.0 | 17 | 2.8 | 2.7 | 2.9 | 5.159 | 0.008 | C19+ < C19- p=0.01 |
| aparc-DKTatlas_lh_volume_superiorfrontal | 25 | 26990.0 | 23909.5 | 28520.5 | 34 | 25332.0 | 24177.0 | 27151.8 | 17 | 22980.0 | 22287.0 | 26677.0 | 3.518 | 0.035 | C19+ < HC p=0.049 |
| aparc-DKTatlas_rh_area_transversetemporal | 25 | 313.0 | 295.5 | 333.5 | 34 | 321.5 | 300.0 | 351.3 | 17 | 329.0 | 293.5 | 359.5 | 3.655 | 0.031 | C19- > HC p=0.027 |
| aparc-DKTatlas_rh_thickness_middletemporal | 25 | 3.0 | 3.0 | 3.1 | 34 | 2.9 | 2.8 | 3.0 | 17 | 3.0 | 2.8 | 3.1 | 3.468 | 0.036 | C19- < HC p=0.032 |
| aparc-DKTatlas_rh_thickness_supramarginal | 25 | 2.9 | 2.8 | 3.0 | 34 | 2.8 | 2.7 | 2.9 | 17 | 2.8 | 2.6 | 2.9 | 3.933 | 0.024 | C19- < HC p=0.031 |
| aparc-DKTatlas_rh_volume_isthmuscingulate | 25 | 2685.0 | 2550.0 | 2947.0 | 34 | 2476.0 | 2224.0 | 2765.0 | 17 | 2462.0 | 2266.0 | 2653.0 | 4.374 | 0.016 | C19- < HC p=0.02 |
| T2_FLAIR_WMH_volume | 25 | 1457.0 | 639.5 | 2726.0 | 34 | 1861.0 | 1018.3 | 2891.0 | 16 | 3765.0 | 2299.0 | 7556.3 | 3.722 | 0.029 | C19+ > HC p=0.027 |
| T2_FLAIR_PVWMH_volume | 25 | 1305.0 | 522.0 | 2323.5 | 34 | 1484.5 | 802.8 | 2417.0 | 16 | 2862.5 | 1839.3 | 6405.5 | 4.921 | 0.01 | C19+ > HC p=0.009; C19+ > C19- p=0.044 |
| dMRI_TBSS_MD_Sagittal_stratum_R | 25 | 813.0 | 787.5 | 829.0 | 34 | 822.0 | 789.3 | 842.3 | 17 | 872.0 | 846.5 | 888.5 | 6.082 | 0.004 | C19+>HC p=0.003; C19+>C19- p=0.037 |
| dMRI_TBSS_MD_NAMW_Sagittal_stratum_R | 25 | 813.0 | 786.5 | 829.0 | 34 | 822.0 | 789.3 | 842.3 | 16 | 870.0 | 843.8 | 891.3 | 6.728 | 0.002 | C19+>HC p=0.002; C19+>C19- p=0.019 |
| SWI_T2star_r_thalamus | 23 | 42.4 | 40.0 | 45.2 | 34 | 45.0 | 43.2 | 46.0 | 17 | 42.1 | 40.7 | 43.3 | 4.396 | 0.016 | C19->HC p=0.022 |

The table reports results for the IDPs which showed a significant group difference and at least one significant post-hoc pairwise difference. Median and interquartile range of raw data are presented in the table for ease of interpretation. P-values were derived from one-way ANOVA comparison of gaussianised deconfounded data and subsequent pairwise comparisons, Bonferroni corrected. Supplementary figure S2 shows the anatomical location of the brain areas involved.

### Supplementary figures

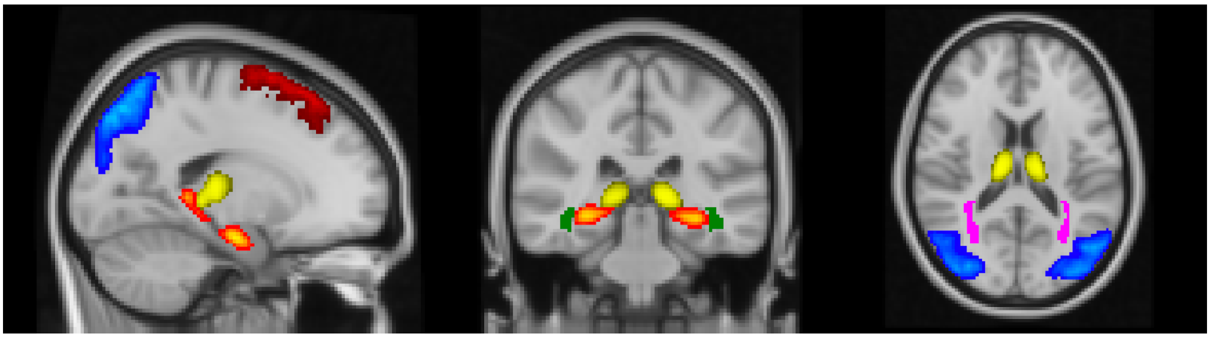

**Figure S1**. Brain areas showing a significant difference between COVID-19 patients and controls in one or more MRI modality (see Table S3). Red-Yellow = hippocampus; Blue-light blue = lateral occipital cortex (superior division); Red = Superior frontal gyrus; Green = sagittal stratum; Magenta = posterior thalamic radiation; Yellow = thalamus.

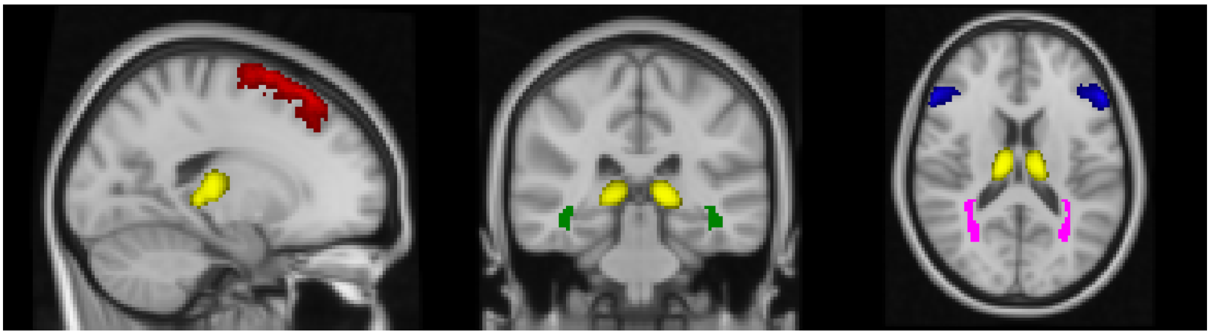

**Figure S2**. Brain areas showing a significant difference across severity groups (COVID-19 who received organ support, COVID-19 patients who did not receive organ support and controls) in one or more MRI modality (see Table S4). Red = Superior frontal gyrus; Blue = Inferior frontal gyrus (pars triangularis); Green = sagittal stratum; Magenta = posterior thalamic radiation; Yellow = thalamus.
